# Association between environmental levels of PM_2.5_ and mortality from SARS-CoV-2 in inhabitants of Mexico City

**DOI:** 10.1101/2022.05.08.22274818

**Authors:** Moreno-Santillan Armando Alberto, Rivas-Ruiz Rodolfo, Fuentes-Valdivieso Rocio, Roy-Garcia Ivonne Anali

**Author notes:** **Corresponding Author** Dr. Rodolfo Rivas Ruiz.

## Abstract

**Objective:** To evaluate the association between exposure to environmental levels of PM_2.5_ and mortality from SARS-CoV-2 in inhabitants of Mexico City.

**Material and methods:** A secondary analysis with the total number of deaths from COVID-19 in residents of Mexico City as well as 25 municipalities in the interior of the Republic was carried out. Environmental levels of PM_2.5_ were between 2018 and 2021. Bivariate analysis and multivariate logistic regression were performed.

**Results:** A total of 1,083,175 cases of COVID-19 were included, with 57,384 deaths (5.3%), of which 30,561 were in residents with exposure to more than 20 μg/m^3^ of PM_2.5_ (OR 1.27, CI95%: 1.25 to 1.29). When performing the multivariate analysis, an OR of 1.39 (CI95%: 1.36 to 1.43) was observed.

**Conclusions:** Chronic exposure to elevated levels of PM_2.5_ is associated with increased death risk from COVID-19.

## INTRODUCTION

In December 2019, the Chinese province of Wuhan became the epicenter of an unknown etiology pneumonia, that as the days passed, started to increase the global attention and concern. On January the 7^th^ of 2020, Chinese health authorities identified the causative agent as an RNA virus that would be known as SARS-CoV-2, which caused the COVID-19 disease.^1^ At the time of writing of this manuscript, the disease has caused more than 490,000,000 cases of contagion and more than 6,100,000 deaths worldwide. In addition, more than 5,700,000 cases and 320,000 deaths have been reported in Mexico. ^2,3^

One of the epidemiological aspects that has drawn attention to the infections and deaths caused by the COVID-19 is its variable behavior between different regions, which could be explained in part by socioeconomic, demographic, geographical, and climatic conditions. ^4,5^ Pansini and Fornacca published one out of the first studies documenting the proportional relationship between average annual levels of particulates less than 2.5 and 10 microns (PM_2.5_ and PM_10_), nitrogen dioxide (NO_2_), ozone (O_3_) and carbon monoxide (CO) in China, Italy, and the United States of America (USA) with the number of SARS-CoV-2 infections. ^6^ This relationship has been confirmed by subsequent studies that have shown that exposure to high levels of pollution may increase the vulnerability of the population to the effects of COVID-19.^7^ This is based on the fact that chronic exposure to high concentrations of PM_2.5_ has been associated with the development and/or exacerbation of respiratory diseases such as asthma, bronchitis and chronic obstructive pulmonary disease (COPD).^8,9^ Because of their size and aerodynamic capacity, PM_2.5_ are able to evade the filtration system of the respiratory system, reach the alveoli and accumulate by diffusion, which generates an inflammatory and thrombogenic response, as well as increasing the number of angiotensin-2 converting enzyme (ACE-2) receptors in the respiratory epithelium, which is the same receptor the protein S blinds to the SARS-CoV-2. ^7, 10, 11, 12^ In addition, the relation between the PM to COVID-19 mortality could have other explanations as these particles are composed of a mixture of organic and inorganic substances such as sulfates, nitrates, ammonium, sodium chloride, coal, dust, and water, which could serve as a vector increasing air transmissibility.^13,14^

The aim of this study is researching the probable association between chronic exposure to elevated environmental levels of PM_2.5_ (greater than 20 μg/m^3^) and COVID-19 mortality in Mexico City inhabitants. In addition, as a secondary objective, studying this association with other pollutants such as PM_10_, NO_2_ and O_3_ was proposed.

## MATERIAL AND METHODS

With cut-off date of August 27, 2021, a transversal secondary analysis of the open database of the National Epidemiological Surveillance System of the General Directorate of Epidemiology of the Mexican Ministry of Health was conducted, where the total number of confirmed cases and deaths by COVID-19 in residents of the City Halls of Mexico City were included. In addition, residents of the capitals of the states of Aguascalientes, Chiapas, Chihuahua, Durango, Oaxaca, Queretaro and Yucatan were included; as well as residents of the municipalities of Abasolo, Celaya, Guanajuato, Irapuato, Leon, Salamanca, San Luis de la Paz, San Miguel de Allende and Silao in the State of Guanajuato; as well as Atononilco, Atitalaquia, Pachuca, Tizayuca, Tepeji del Rio, Tula, Huichapan, Tulancingo and Tepeapulco in the State of Hidalgo. On the number of cases and deaths by COVID-19, information was obtained, as well as on the main demographic variables and population comorbidities. Subsequently, the environmental levels of PM_2.5_ were recorded, as well as PM_10_, NO_2_ and O_3_ corresponding to the subject municipalities or mayors, which was obtained from the reports of the Automatic Environmental Monitoring Networks of the National Air Quality Information System from January 1, 2018 to August 27, 2021. Cases of immigrants and those in which SARS-CoV-2 infection could not be confirmed were excluded, such as studies invalidated by the laboratory, cases without confirmatory evidence, suspicious cases or with negative test, as well as the reports of mayors or municipalities with measurements of absent or incomplete environmental pollutants, for which the criterion of the Mexican Official Standard NOM-025-SSA1-2014 was taken. In this case, it is established that there must be at least three quarters with at least 75% of the data.^15^

For univariate analysis, quantitative variables are presented in measures of central tendency and dispersion according to their type of distribution, and qualitative variables are presented in percentages. For the bivariate statistical analysis, the U Mann Whitney and Squared Chi tests was used. To establish a comparison between groups, average exposure to more than 20 μg/m^3^ for PM_2.5_, average exposure to more than 35 μg/m^3^ for PM_10_, average exposure to more than 10 ppb for NO_2_, and average exposure to more than 45 ppb for O_3_ were established as cutoff points. To adjust the main confounders, a multiple logistic regression model was performed considering the main comorbidities associated with mortality from COVID-19 such as diabetes, arterial hypertension, obesity, asthma, cardiovascular disease (CVD), chronic renal disease (CRD), chronic obstructive pulmonary disease (COPD) and immunosuppression. For all tests, the values of p <=0.05 were considered as statistically significant. RStudio statistical software version 4.1.0 © 2009-2021 was used.

The present study was evaluated and approved by the Local Research Committee of the High Specialty Medical Unit “Luis Castelazo Ayala” of the Mexican Social Security Institute under number R-2021-3606-011. Since it was carried out based on reviewed documentary files, and in accordance with the General Health Law on Research, it was classified as presenting a “lower risk to the minimum”.

## RESULTS

1,083,175 confirmed cases of COVID-19 were studied in the selected regions, of which 759,087 people (70.1%) were residents of Mexico City, and 324,039 people (29.9%) were residents of the country’s inland. Of the total population, 57,384 deaths were recorded (5.3%), which represents a mortality rate of 310.7 per 100,000 inhabitants and a case fatality rate of 0.05%. The general characteristics of the study population are presented in Table 1. Furthermore, the distribution of confirmed cases, prevalence (per 100,000 inhabitants), deaths, mortality rate (per 100,000 inhabitants) and annual average of PM_2.5_ per Mayor or Municipality of study is presented in Table 2.

**Table 1.** Baseline characteristics of the study population (N=1,083,175).

| Variable | $PM_{2.5} < 20 \mu g/m^3$ | $PM_{2.5} > 20 \mu g/m^3$ | p |
| --- | --- | --- | --- |
| Age in years, median (IQR) | 38 (27, 52) | 40 (28, 53) | < 0.01* |
| Female sex, N (%) | 274,748 (25.4) | 252,477 (23.3) | < 0.01** |
| Comorbidities, N (%) |  |  |  |
| • Arterial hypertension | 70,427 (6.5) | 66,961 (6.2) | < 0.01** |
| • Obesity | 57,167 (5.3) | 57,875 (5.3) | < 0.01** |
| • Diabetes | 52,480 (4.8) | 51,130 (4.7) | < 0.01** |
| • Smoking | 45,761 (4.2) | 46,013 (4.2) | < 0.01** |
| • Asthma | 10,566 (1.0) | 10,532 (1.0) | < 0.01** |
| • Cardiovascular disease | 5,828 (0.5) | 6,361 (0.6) | < 0.01** |
| • Chronic renal disease | 5,053 (0.5) | 5,183 (0.5) | < 0.01** |
| • Chronic obstructive pulmonary disease | 3,894 (0.4) | 4,342 (0.4) | < 0.01** |
| • Immunosuppression | 3,275 (0.3) | 3,259 (0.3) | < 0.01** |
| Deaths, N (%) | 26,823 (2.5) | 30,561 (2.8) | < 0.01** |
| Survivors, N (%) | 540,765 (49.9) | 485,026 (44.8) | < 0.01** |

\*U Mann Whitney \*\* Squared Chi. IQR = Interquartile range.

**Table 2.** Distribution of cases and deaths from COVID-19 and average exposure to PM_2.5_ levels.

| State | Municipality or mayor | Confirmed cases * | Prevalence | COVID-19 deaths | Mortality rate | PM<sub>2.5</sub> levels ** |
| --- | --- | --- | --- | --- | --- | --- |
| Mexico City | Alvaro Obregon | 12,7394 | 16,781.4 | 3,137 | 413.2 | 17.4 |
| Mexico City | Azcapotzalco | 45,424 | 10,509.8 | 2,554 | 590.9 | 23.5 |
| Mexico City | Benito Juarez | 30,459 | 7,015.7 | 1,373 | 316.2 | 20.1 |
| Mexico City | Coyoacan | 48,684 | 7,923.2 | 2,546 | 414.4 | 19.7 |
| Mexico City | Cuajimalpa | 16,013 | 7,356 | 556 | 255.4 | 17.5 |
| Mexico City | Cuauhtemoc | 45,448 | 8,325.6 | 2,506 | 459.1 | 24 |
| Mexico City | Gustavo A. Madero | 101,198 | 8,624.7 | 5,792 | 493.6 | 23.7 |
| Mexico City | Iztapalapa | 140,013 | 7,628.1 | 7,491 | 408.1 | 23.2 |
| Mexico City | Miguel Hidalgo | 30,270 | 7,303.3 | 1,333 | 321.6 | 25 |
| Mexico City | Tlalpan | 85,176 | 12,169.3 | 2,013 | 287.6 | 18.9 |
| Mexico City | Venustiano Carranza | 41,809 | 9,422.7 | 2,144 | 483.2 | 23.8 |
| Mexico City | Xochimilco | 47,208 | 10,676.2 | 1,331 | 301.0 | 16.7 |
| Aguascalientes | Aguascalientes | 25,661 | 2,704.0 | 2,256 | 237.7 | 11.1 |
| Chiapas | Tuxtla Gutierrez | 8,085 | 1,338.3 | 730 | 120.8 | 17 |
| Chihuahua | Chihuahua | 20,028 | 2,135.9 | 1,941 | 207.0 | 13.6 |
| Durango | Durango | 24,721 | 3,589.5 | 1,242 | 180.3 | 16.45 |
| Guanajuato | Abasolo | 748 | 812.7 | 98 | 106.5 | 14.6 |
| Guanajuato | Celaya | 13,595 | 2,608.6 | 958 | 183.8 | 19.8 |
| Guanajuato | Guanajuato | 6,176 | 3,175.3 | 374 | 192.3 | 12.6 |
| Guanajuato | Irapuato | 15,248 | 2,571.5 | 1,224 | 206.4 | 19.7 |
| Guanajuato | Leon | 50,651 | 2,942.7 | 4,167 | 242.1 | 20.6 |
| Guanajuato | Salamanca | 7,471 | 2,732.5 | 604 | 220.9 | 21.9 |
| Guanajuato | San Luis de la Paz | 2,744 | 2,134.8 | 214 | 166.5 | 11.5 |
| Guanajuato | San M. de Allende | 3,558 | 2,037.6 | 230 | 131.7 | 13.1 |
| Guanajuato | Silao | 3,763 | 1,848.6 | 397 | 195.0 | 25.1 |
| Hidalgo | Atotonilco de Tula | 880 | 1,408.7 | 126 | 201.7 | 32.4 |
| Hidalgo | Atitalaquia | 649 | 2,058.7 | 119 | 377.5 | 23 |
| Hidalgo | Pachuca | 11,291 | 3,592.1 | 1,277 | 406.3 | 21.7 |
| Hidalgo | Tizayuca | 4,091 | 2,430.7 | 457 | 271.5 | 21.7 |
| Hidalgo | Tepeji del Rio | 2,130 | 2,352.4 | 261 | 288.3 | 20.9 |
| Hidalgo | Tula de Allende | 2,718 | 2,361.3 | 402 | 349.2 | 19.3 |
| Hidalgo | Huichapan | 798 | 1,682.7 | 91 | 191.9 | 15.5 |
| Hidalgo | Tulancingo | 4,098 | 2,433.9 | 507 | 301.1 | 14.6 |
| Hidalgo | Tepeapulco | 2,001 | 3,557.6 | 206 | 366.3 | 14.2 |
| Oaxaca | Oaxaca | 16,846 | 6,217.3 | 835 | 308.2 | 11.9 |
| Queretaro | Queretaro | 57,996 | 5,524.6 | 3,169 | 301.9 | 12.3 |
| Yucatan | Merida | 38,092 | 3,827.8 | 2,735 | 274.8 | 12.8 |

\* National Epidemiological Surveillance System \*\* Annual average levels between 2018-2021

The result of the bivariate analysis (using Pearson’s Squared Chi), which establishes the association between mortality caused by COVID-19 and the average exposure to more than 20 μg/m^3^ of PM _2.5_, more than 35 μg/m^3^ of PM_10_, more than 10ppb of NO_2_, and more than 45 ppb of O_3_ in the study population, as well as the main comorbidities associated with COVID-19 mortality are presented in Table 3. In addition, the Population Attributable Risk (PAR) values, expressed in percentage, are presented.

**Table 3.** Risk of death from COVID-19 according to exposure to environmental pollutants and the presence of comorbidities.

|  |  | **Deaths**<br>N=57,384 (%) | **Survivors**<br>N=1,025,742 (%) | **OR\*** | **95% IC** | **PAR (%)** |
| --- | --- | --- | --- | --- | --- | --- |
| **Environmental pollutant** | > 20 µg/m<sup>3</sup> PM<sub>2.5</sub> | 30,561 (53.2) | 485,026 (47.2) | 1.27 | 1.25 - 1.29 | 14.7 |
|  | > 35 µg/m<sup>3</sup> PM<sub>10</sub> | 38,151 (66.5) | 596,079 (58.1) | 1.81 | 1.78 - 1.83 | 21.1 |
|  | > 10 ppb NO<sub>2</sub> | 45,334 (79) | 854,257 (83.2) | 1.32 | 1.29 - 1.34 | 13.9 |
|  | > 45 ppb O<sub>3</sub> | 30,230 (52.6) | 680,182 (66.3) | 0.57 | 0.52 - 0.58 | - |
| **Comorbidities** | Diabetes | 20,324 (35.4) | 83,286 (8.1) | 6.21 | 6.09 - 6.32 | 28.4 |
|  | Arterial hypertension | 24,985 (43.5) | 112,403 (10.9) | 6.27 | 6.16 - 6.38 | 34.8 |
|  | Obesity | 12,059 (21.1) | 102,983 (10.1) | 2.38 | 2.33 - 2.43 | 11.6 |
|  | Cardiovascular disease | 2,699 (4.7) | 9,480 (0.9) | 5.29 | 5.06 - 5.52 | 3.5 |
|  | Chronic renal disease | 3,656 (6.4) | 6,580 (0.6) | 10.5 | 10.1 - 11.0 | 5.4 |
|  | COPD | 2,568 (4.4) | 5,668 (0.5) | 8.43 | 8.04 - 8.84 | 3.7 |
|  | Immunosuppression | 1,150 (2.1) | 5,384 (0.5) | 3.88 | 3.63 - 4.13 | 1.4 |
| **Male sex** |  | 33,493 (58.4) | 463,406 (45.2) | 1.88 | 1.84 - 1.91 | 28.5 |

COPD = Chronic obstructive pulmonary disease. PAR = Population Attributable Risk \* Squared Chi.

For the multivariate analysis, a logistic regression model was performed, which included the pollutants studied and the main comorbidities associated with COVID-19 (Table 4 and Figure 1). COVID-19 mortality was found to have an adjusted OR of 1.39 (95% CI: 1.36 to 1.43) when exposure was greater than 20 μg/m^3^ of PM2.5, as well as OR of 1.15 (95% CI 1.12 to 1.18) for exposure greater than 35 μg/m^3^ of PM_10_, of 1.02 (95% CI 1.02 to 1.07) for exposure of more than 10 ppb of NO_2_, and OR of 0.56 (95% CI 0.55 to 0.57) for exposure of more than 45 ppb of O_3_.

**Table 4.** Multivariate model with the adjustment of contaminants and the main comorbidities associated with COVID-19.

|  | COVID-19 deaths (n= 57,384) |  |  |  |
| --- | --- | --- | --- | --- |
|  | B | OR | 95% IC | p |
| > 20 $\mu\text{g}/\text{m}^3$ PM<sub>2.5</sub> | 0.33 | 1.39 | 1.36 - 1.43 | < 0.01 |
| > 35 $\mu\text{g}/\text{m}^3$ PM<sub>10</sub> | 0.14 | 1.15 | 1.12 - 1.18 | < 0.01 |
| > 10 ppb NO<sub>2</sub> | 0.04 | 1.04 | 1.02 - 1.07 | 0.002 |
| > 45 ppb O<sub>3</sub> | -0.56 | 0.56 | 0.55 - 0.57 | < 0.01 |
| Diabetes | 1.08 | 2.95 | 2.89 - 3.02 | < 0.01 |
| Arterial hypertension | 1.17 | 3.22 | 3.15 - 3.29 | < 0.01 |
| Obesity | 0.38 | 1.47 | 1.43 - 1.51 | < 0.01 |
| Cardiovascular disease | 0.51 | 1.68 | 1.59 - 1.77 | < 0.01 |
| Chronic renal disease | 1.16 | 3.20 | 3.05 - 3.37 | < 0.01 |
| Chronic obstructive pulmonary disease | 1.23 | 3.45 | 3.26 - 3.65 | < 0.01 |
| Immunosuppression | 0.67 | 1.95 | 1.80 - 2.11 | < 0.01 |

Nagelkerke $R^2 = 0.16$ , $p < 0.001$ .

**Figure 1.**
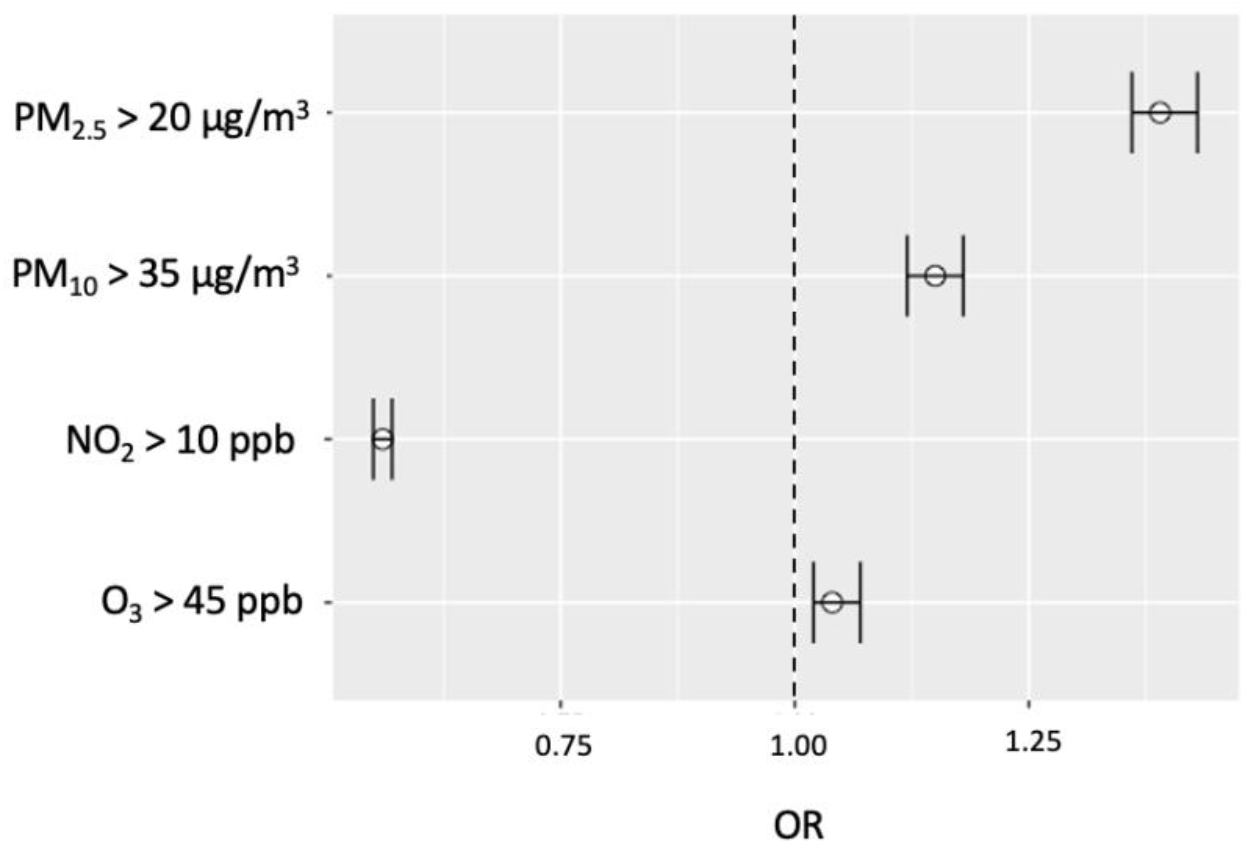
Graphic representation of the multivariate analysis where the association between environmental contaminants and risk of death from COVID-19 is established.

## DISCUSSION

It has been observed that COVID-19 mortality has a relationship with the presence of comorbidities such as arterial hypertension, diabetes, heart disease, COPD, renal disease, or immunosuppression. However, there are other factors as environmental pollutants, which can play a relevant role. In this sense, and based on the results of our study, we observed that chronic exposure to high levels of PM_2.5_, PM_10_ and NO_2_ represents an increased risk of death from SARS-CoV-2 infection, which when expressed in terms of the Attributable Population Risk (PAR) represented a 14.7% for PM_2.5_, 21.1% for PM_10_, and 13.9% for NO_2_. It is worth mentioning that the PAR represents the disease incidence proportion that would be avoided in the general population if exposure to the risk factor were eliminated. In this case, it was related to the environmental pollutants. Unlike the ratio of possibilities (OR) and relative risk (RR), the PAR takes into account the number of individuals exposed in the population. The usefulness of the PAR calculation is that it can be used to plan public health programmes in which specific disease prevention strategies need to be identified. In other words, based on our study, it could serve to identify the population impact of COVID-19 deaths when implementing environmental measures (PM_2.5_, PM_10_ and NO_2_) aimed at reducing the levels of environmental pollutants below the limits studied.^16.17^

When analyzing the findings of the present study, we observed that they are consistent with those documented in other scientific reports that, although they are based on different methodologies, they have confirmed the relationship between death by COVID-19 and exposure to high levels of environmental pollutants. Frontera’s publication was one of the first ones, who in May 2020 observed a greater number of cases of death by COVID-19 in the most polluted regions of northern Italy, being up to twice as many compared to less polluted areas.^18^ On their behalf, Wu and collaborators at Johns Hopkins University conducted quantitative research on the role of PM_2.5_ and COVID-19 mortality, and reported that by the increase in 1 μg/m^3^ of PM_2.5_, the risk of death by SARS-CoV-2, increase in 8% ^19^, being this study one of the most important, since it included the study of 3,000 US counties (representative of 98% of the population) and whose results give rise to the present research. In Italy, Coker also observed that the 1 μg/m^3^ increase in PM_2.5_ was associated with 9% (95% CI: 6-12%) increased risk of death from COVID-19.^20^ These results are below the level of risk found in our study, which could be due to the statistical methodology and the higher levels of PM_2.5_ of our study regions. In Latin America, Vasquez-Apestegui and collaborators were the first to study the population exposure to PM2.5 between 2010 and 2016 in 24 Lima districts, and to establish their relationship with COVID-19 they applied a multivariate regression model, noting a significant association between cases and deaths with high exposure to PM_2.5_.^21^ In Mexico, Cabrera-Cano and collaborators conducted an ecological study in 25 cities, in which they found a non-significant association between levels of PM_2.5_ and mortality by COVID-19, which differs from our results. In this study, only the pollutant reports of five months of the year 2020 were considered.^22^ Due to the fact that, we could present the discrepancy.

Regarding the study of PM_10_, Márques and collaborators studied in Barcelona the effect of high chronic exposure to this pollutant, in which they observed a higher death risk caused by COVID-19 (OR 2.37, 95% CI 1.71-3.32) than some comorbidities such as asthma, obesity, diabetes or COPD, concluding that per 1 µg/m^3^ increase in PM_10_, the death risk caused by COVID-19 increases by 2.68% (95% CI 0.53%-5.58%). ^23^ Additionally, when studying 55 Italian provinces, Coccia observed a higher number of COVID-19 infections in regions that exceeded the PM_10._ limit levels by more than 100 days. ^24^ By measuring global exposure to fine atmospheric particles by satellite, Pozzer estimated that environmental pollution contributes 15% to COVID-19 mortality. ^25^ Although the afore mentioned studies were carried out under different methodologies, their results are compatible with the findings of our study, in which we observed a 15% increase in death risk caused by COVID-19 in regions whose exposure to PM_10_ is equal to or greater than 35 μg/m^3^.

In the case of studies that include NO_2_, in China, Zhu observed that due to the increase of every 10 μg/m^3^ of NO_2_, there was a 6.94% increase of COVID-19 daily cases.^26^ In addition, in the United States, Liang and collaborators reported that due to the increase in the inter-quartile range of NO_2_ (4.6 ppb), there was an 11.2% increase (95% CI 3.4 to 19.5%) in mortality due to COVID-19.^27^ In this way, it coincides with our study, in which we observed a PAR of 13.9% and a discrete increase of 4% in the death risk caused by COVID-19 with chronic exposure to more than 10 ppb of NO_2_. In their respective studies, Zoran and Bashir documented results different from ours, finding no association between the death risk and the exposure to high levels of NO_2_.^28,29^

Few studies have explored the effect of O_3_ on mortality caused by COVID-19. One of them was that of Ayoub, who reported that due to the increase in 1 unit of ozone, there were 4.4% more deaths caused by SARS-CoV-2. ^30^ In New York, Adhikari and collaborators observed that acute exposure to high ozone concentrations, along with weather variables such as wind speed, temperature, and humidity, was associated with more cases of COVID-19.^31^ These results differ from our study findings, where not only we could not verify the association between ozone exposure and mortality caused by COVID-19, but we also observed an apparent protective effect (OR 0.56, 95%CI; 0.55 to 0.57), which should be taken with reservation, since the methodological design of our study did not include the evaluation of atmospheric and environmental variables.

One of the challenges in developing the methodological design of the present study was the choice of cut-off points for PM concentrations as the WHO, in its Guide to Global Air Quality 2021 states that the recommended levels for avoiding health risks are 5 μg/m^3^ for PM_2.5_, and 15 μg/m^3^ for PM_10_. However, these references cannot be applied in our country context, since no region studied has registered equal or lower levels than those recommended, which could be interpreted as the need to establish more realistic cut-off points according to the current context, or that national environmental policies need to be thoroughly reviewed and improved.^8^

The pandemic is not over, so our results have a limitation inherent in its final course, as well as other variables that may affect mortality by COVID-19 and that were not included in the study, such as the socioeconomic stratum, population mobility, vaccination coverage, the effect of seasons and the temperature. Another limitation of the study was the measurement of pollutants levels, since not all are consistent and homogeneous in the different monitoring stations, so four Mexico City Mayors could not be included (Iztacalco, Magdalena Contreras, Milpa Alta and Tlahuac). For the same reason, it was not possible to explore other pollutants such as carbon monoxide, nitric oxide or sulphur dioxide. However, the measurements of PM_2.5_, PM_10_, NO_2_ and O_3_ that were included in the study, filled the national Air Quality Standards established in the “Official Mexican Environmental Health Standard. Permissible limit values for the concentration of suspended particles PM_10_ and PM_2.5_ in air and criteria for their assessment”. ^15^ Therefore, more studies will be needed to help differentiate the role of confounders and environmental pollutants as risk factors in COVID-19 mortality.

## CONCLUSIONS

Chronic exposure to elevated levels of PM_2.5,_ as well as PM_10_ and NO_2_ is associated with increased death risk caused by COVID-19 in Mexico City residents.

## Data Availability

All data produced are available online at:
https://datos.gob.mx/busca/dataset/informacion-referente-a-casos-covid-19-en-mexico
https://sinaica.inecc.gob.mx/

https://datos.gob.mx/busca/dataset/informacion-referente-a-casos-covid-19-en-mexico

https://sinaica.inecc.gob.mx/

## Acknowledgment

To the National Polytechnic Institute, Mexico.

## Notes

### Competing Interest Statement

The authors have declared no competing interest.

### Funding Statement

This study did not receive any funding

### Author Declarations

https://datos.gob.mx/busca/dataset/informacion-referente-a-casos-covid-19-en-mexico https://sinaica.inecc.gob.mx/

## REFERENCES

1. Wang C, Horby PW, Hayden FG, Gao GF. A novel coronavirus outbreak of global health concern. The Lancet. 2020;395(10223). doi:10.1016/S0140-6736(20)30185-9

2. Yesudhas D, Srivastava A, Gromiha MM. COVID-19 outbreak: history, mechanism, transmission, structural studies and therapeutics. Infection. 2021;49(2). doi:10.1007/s15010-020-01516-2

3. Center for Systems Science and Engineering. COVID-19 Map -Johns Hopkins Coronavirus Resource Center. Johns Hopkins Coronavirus Resource Center. Published online 2021.

4. Bellali H, Chtioui N, Chahed M. Factors associated with country-variation in COVID-19 morbidity and mortality worldwide: An observational geographic study COVID-19 morbidity and mortality country-variation. medRxiv. Published online 2020. doi:10.1101/2020.05.27.20114280

5. Biktasheva I v. Role of a habitat’s air humidity in Covid-19 mortality. Science of the Total Environment. 2020;736. doi:10.1016/j.scitotenv.2020.138763

6. Pansini R, Fornacca D. Initial evidence of higher morbidity and mortality due to SARS-CoV-2 in regions with lower air quality. medRxiv. Published online 2020.

7. Amnuaylojaroen T, Parasin N. The Association Between COVID-19, Air Pollution, and Climate Change. Frontiers in Public Health. 2021;9. doi:10.3389/fpubh.2021.662499

8. Health Organization W. Ambient air pollution: a global assessment of exposure and burden of disease. Clean Air Journal. 2016;26(2). doi:10.17159/2410-972x/2016/v26n2a4

9. Shin S, Bai L, Burnett RT, et al. Air pollution as a risk factor for incident chronic obstructive pulmonary disease and Asthma: A 15-year population-based cohort study. American Journal of Respiratory and Critical Care Medicine. 2021;203(9). doi:10.1164/rccm.201909-1744OC

10. Cole MA, Ozgen C, Strobl E. Air Pollution Exposure and Covid-19 in Dutch Municipalities. Environmental and Resource Economics. 2020;76(4). doi:10.1007/s10640-020-00491-4

11. Xing YF, Xu YH, Shi MH, Lian YX. The impact of PM2.5 on the human respiratory system. Journal of Thoracic Disease. 2016;8(1). doi:10.3978/j.issn.2072-1439.2016.01.19

12. Beyerstedt S, Casaro EB, Rangel ÉB. COVID-19: angiotensin-converting enzyme 2 (ACE2) expression and tissue susceptibility to SARS-CoV-2 infection. European Journal of Clinical Microbiology and Infectious Diseases. 2021;40(5). doi:10.1007/s10096-020-04138-6

13. Travaglio M, Yu Y, Popovic R, Selley L, Leal NS, Martins LM. Links between air pollution and COVID-19 in England. Environmental Pollution. 2021;268. doi:10.1016/j.envpol.2020.115859

14. Landrigan PJ, Fuller R, Acosta NJR, et al. The Lancet Commission on pollution and health. The Lancet. 2018;391(10119). doi:10.1016/S0140-6736(17)32345-0

15. Secretaría de Salud. NORMA Oficial Mexicana NOM-025-SSA1-2014, Salud Ambiental. Valores Límite Permisibles Para La Concentración de Partículas Suspendidas PM10 y PM2.5 En El Aire Ambiente y Criterios Para Su Evaluación.; 2014.

16. Fajardo-Gutiérrez A. Medición en epidemiología: prevalencia, incidencia, riesgo, medidas de impacto. Revista Alergia Mexico. 2017;64(1). doi:10.29262/ram.v64i1.252

17. Javier Nieto García F, Peruga Urrea A. Riesgo Atribuible: Sus Formas, Usos e Interpretación. Gaceta Sanitaria. 1990;4(18). doi:10.1016/S0213-9111(90)71007-2

18. Frontera A, Cianfanelli L, Vlachos K, Landoni G, Cremona G. Severe air pollution links to higher mortality in COVID-19 patients: The “double-hit” hypothesis. Journal of Infection. 2020;81(2). doi:10.1016/j.jinf.2020.05.031

19. Wu X, Nethery RC, Sabath BM, Braun D, Dominici F. Exposure to air pollution and COVID-19 mortality in the United States. ISEE Conference Abstracts. 2020;2020(1). doi:10.1289/isee.2020.virtual.o-os-638

20. Coker ES, Cavalli L, Fabrizi E, et al. The Effects of Air Pollution on COVID-19 Related Mortality in Northern Italy. Environmental and Resource Economics. 2020;76(4). doi:10.1007/s10640-020-00486-1

21. Vasquez-Apestegui B v., Parras-Garrido E, Tapia V, et al. Association between air pollution in Lima and the high incidence of COVID-19: findings from a post hoc analysis. BMC Public Health. 2021;21(1). doi:10.1186/s12889-021-11232-7

22. Cabrera-Cano ÁA, Cruz-de la Cruz JC, Gloria-Alvarado AB, Álamo-Hernández U, Riojas-Rodríguez H. Asociación entre mortalidad por Covid-19 y contaminación atmosférica en ciudades mexicanas. Salud Publica de Mexico. 2021;63(4). doi:10.21149/12355

23. Marquès M, Correig E, Ibarretxe D, et al. Long-term exposure to PM10 above WHO guidelines exacerbates COVID-19 severity and mortality. Environment International. 2022;158. doi:10.1016/j.envint.2021.106930

24. Coccia M. Factors determining the diffusion of COVID-19 and suggested strategy to prevent future accelerated viral infectivity similar to COVID. Science of the Total Environment. 2020;729. doi:10.1016/j.scitotenv.2020.138474

25. Pozzer A, Dominici F, Haines A, Witt C, Münzel T, Lelieveld J. Regional and global contributions of air pollution to risk of death from COVID-19. Cardiovascular Research. 2020;116(14). doi:10.1093/cvr/cvaa288

26. Zhu Y, Xie J, Huang F, Cao L. Association between short-term exposure to air pollution and COVID-19 infection: Evidence from China. Science of the Total Environment. 2020;727. doi:10.1016/j.scitotenv.2020.138704

27. Liang D, Shi L, Zhao J, et al. Urban Air Pollution May Enhance COVID-19 Case-Fatality and Mortality Rates in the United States. Innovation(China). 2020;1(3). doi:10.1016/j.xinn.2020.100047

28. Zoran MA, Savastru RS, Savastru DM, Tautan MN. Assessing the relationship between ground levels of ozone (O3) and nitrogen dioxide (NO2) with coronavirus (COVID-19) in Milan, Italy. Science of the Total Environment. 2020;740. doi:10.1016/j.scitotenv.2020.140005

29. Bashir MF, Ma BJ, Bilal, et al. Correlation between environmental pollution indicators and COVID-19 pandemic: A brief study in Californian context. Environmental Research. 2020;187. doi:10.1016/j.envres.2020.109652

30. Ayoub Meo S, Adnan Abukhalaf A, Sami W, Hoang TD. Effect of environmental pollution PM2.5, carbon monoxide, and ozone on the incidence and mortality due to SARS-CoV-2 infection in London, United Kingdom. Journal of King Saud University - Science. 2021;33(3). doi:10.1016/j.jksus.2021.101373

31. Adhikari A, Yin J. Short-term effects of Ambient Ozone, PM2.5, and meteorological factors on COVID-19 confirmed cases and deaths in Queens, New York. International Journal of Environmental Research and Public Health. 2020;17(11). doi:10.3390/ijerph17114047

